# Intravascular ultrasound-based external elastic lamina measurements improve vertebral artery assessment over 2-dimensional digital subtraction angiography lumenogram

**DOI:** 10.64898/2026.07.22.26357347

**Authors:** Jordan R. Altman, Krishiv Dhupar, Karthikeyan M. Arcot

## Abstract

Vertebral artery stenosis is a condition amenable to treatment with stenting. To accurately size stents, angiography is the most used imaging technique despite its limitation as a lumenogram with few planes of imagery. The technique’s deficiencies are especially apparent at the vertebral artery origin due to image obstruction by surrounding vessels as well as vessel tortuosity. By contrast, intravascular ultrasound (IVUS) can be used to measure the arterial external elastic lamina (EEL), which is closer to the true vessel size and often leads to larger and more appropriate stent sizes being chosen for more effective treatment.

A retrospective measurement comparison study was conducted on eight patients with vertebral artery stenosis. 2D digital subtraction angiography (2D DSA) and IVUS images were taken along the vertebral artery for each patient, and the blood vessel diameter was measured using the two techniques along a continuous segment of the artery. Our results indicated a statistically significant discrepancy between the diameter measurements (p<0.05), with EEL measurements by IVUS showing a mean percent increase of 32.96%, or 1.19 mm, compared to 2D DSA lumen measurements.

The significant measurement contrast between the two techniques can lead to smaller stent sizes being chosen, increasing the risk of restenosis and stent thrombosis due to malapposition. Sizing stents to the EEL is possible with IVUS imaging and can lead to more reliable treatment due to more secure strut placement on the tunica intima and a more complete expansion of the artery.

## Introduction

Stenosis or occlusion of the vertebral artery can result in reduced cerebral perfusion, causing symptoms referable to the posterior circulation such as vertigo, ataxia, diplopia, and speech disturbance.[1] Stenting is a viable solution for vertebral artery stenosis as it effectively increases luminal area and can improve cerebral perfusion, alleviating associated symptoms.[2] However, arterial structure and tortuosity, as well as adjacent vessels, make it challenging to accurately size and place a stent with angiography, especially at the vertebral artery ostium. 2-dimensional digital subtraction angiography (2D DSA) is constrained by the fact that it is a lumenogram [3] as well as by its limited planes of imaging. Such limitations can create varying luminal measurements depending on the chosen angle of view in stenotic arteries.[4]

Studies have shown that using optical coherence tomography (OCT) guided EEL-based (external elastic lamina) measurements over lumen measurements leads to larger stent sizes being chosen.[5] Using a larger stent size for treating stenosis is often preferred because it promotes better vessel wall apposition and lowers the risk of restenosis by ensuring more complete expansion of the narrowed artery segment.[6]

Intravascular ultrasound (IVUS), like OCT, can measure the EEL diameter, or the true non-diseased diameter, of arteries. In this study, we used IVUS to demonstrate that 2D DSA does not measure the true arterial size accurately, especially for the purpose of sizing stents.

## Materials and Methods

### Patient Selection

All eight patients were referred for symptomatic lesions of the vertebral artery with either recurrent postural symptoms (lightheadedness on standing up) or ischemic strokes in the territory referable to the vertebral artery. Included patients had corresponding 2D DSA and IVUS imaging. No additional patients met the criteria for inclusion. Patient ages ranged from 56 to 85 years old (mean = 72.5 ± 11.7), with one female and seven male patients.

### Data Acquisition

Under fluoroscopic guidance, a Benchmark 071 guide catheter was introduced into the subclavian artery. 2D DSA imaging was performed using a Siemens Artis Q biplane imaging system (Figure 1). A 014 microwire was then navigated into the V3 segment of the vertebral artery. The IVUS catheter was then navigated distal to the stenosis, and once the position was confirmed, imaging was performed (Figure 1) with an automated pullback sled at either 0.5 or 1.0 mm/sec. During pullback acquisition, images were obtained at a rate of 30 frames/sec. The images were analyzed and luminal diameters, degree of stenosis, and length of stenosis were then measured. The IVUS catheter was then used to mark the landing zone for the proximal marker of the stent at or closely adjacent to the ostium of the vertebral artery with a single-shot X-ray. The IVUS catheter was then removed. The appropriately sized stent, as determined by IVUS, was then navigated into the vertebral artery. The proximal marker was positioned at the previously determined point and the stent was deployed. The pusher was removed. The IVUS catheter was again introduced over the wire positioned distal to the stent and an automated pullback was performed. The images were then analyzed, and a post-angioplasty was performed with a non-compliant balloon if necessary.

**Figure 1.**
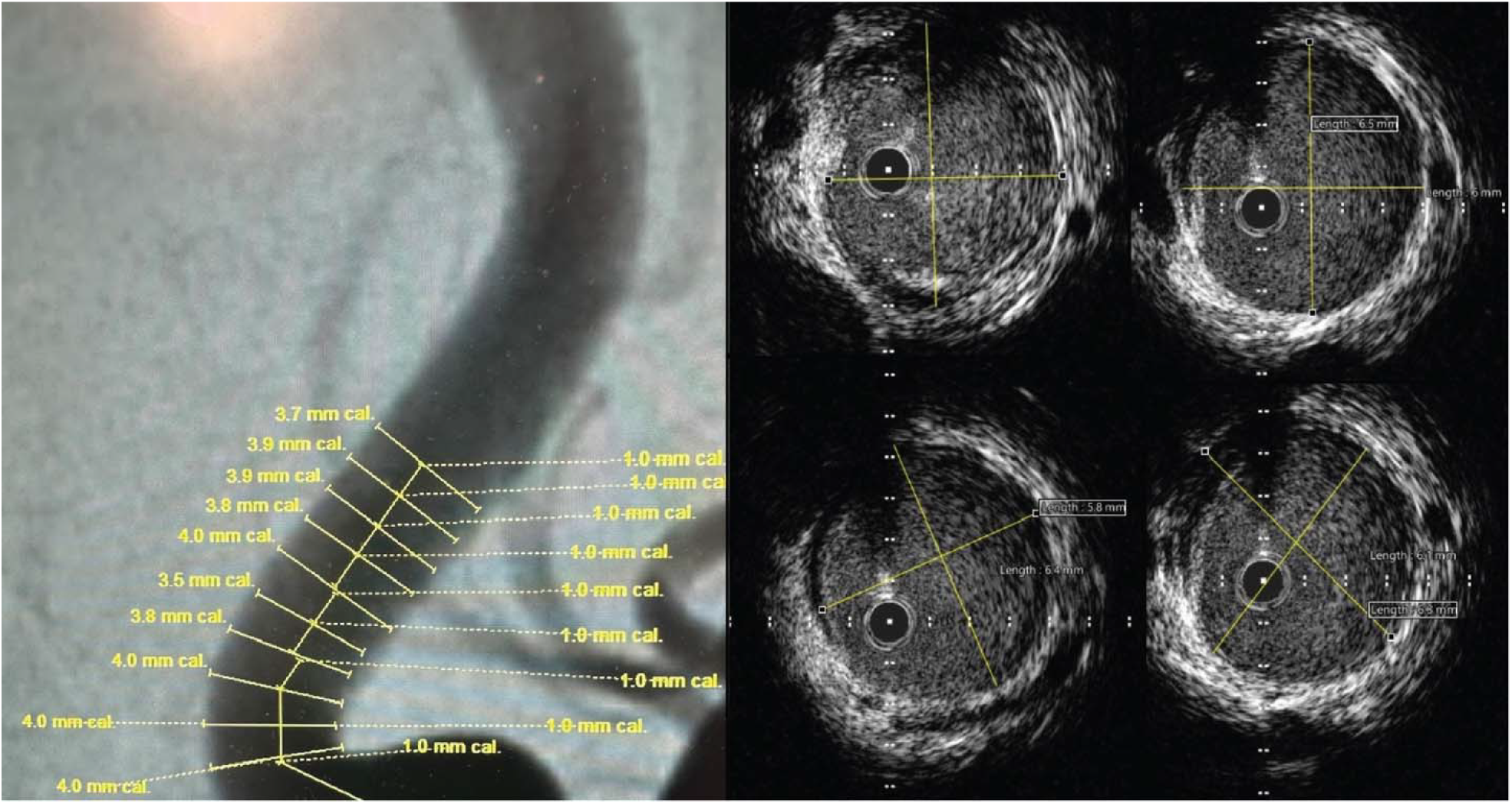
2D Digital Subtraction Angiography (2D DSA) and Intravascular Ultrasound (IVUS). Sample images displaying how measurements of the vertebral artery were taken. Figure 1 (left) shows measurements of the vertebral artery lumen using 2-dimensional digital subtraction angiography images from the Siemens Artis Q biplane imaging system. Figure 1 (right) shows measurements of the external elastic lamina in four separate cross-sections of the vertebral artery using Philips ACIST intravascular ultrasound images.

2D DSA images were then obtained at the level of the stent and of the posterior circulation to ensure that there were no stent abnormalities or distal occlusions.

### Measurement Comparisons

The difference in EEL and lumen diameter was then evaluated in the eight patients with vertebral artery stenosis. We selected 6 to 10 points of interest from each patient, chosen based on the amount of continuous non-stenotic artery available as viewed by both angiogram and IVUS imaging. The length of these segments then determined the intervals chosen between points of interest. The lumen diameter was measured in 2D DSA at a constant window width of 255 and level of 150 to ensure that windowing did not alter vessel measurements between patients. The EEL was then measured at the same 6 to 10 points in each patient using IVUS (Figure 1). All measurements were taken once per point of interest by an interventional neurologist with nine years of experience, blinded to the corresponding values from the other imaging modality.

## Results

Figure 2 presents a comparative analysis of each patient’s mean vertebral artery measurement using IVUS and 2D DSA along the 6 to 10 chosen points. On average, IVUS measurements exceeded 2D DSA measurements by 32.96%, corresponding to a mean difference of 1.19 mm.

**Figure 2.**
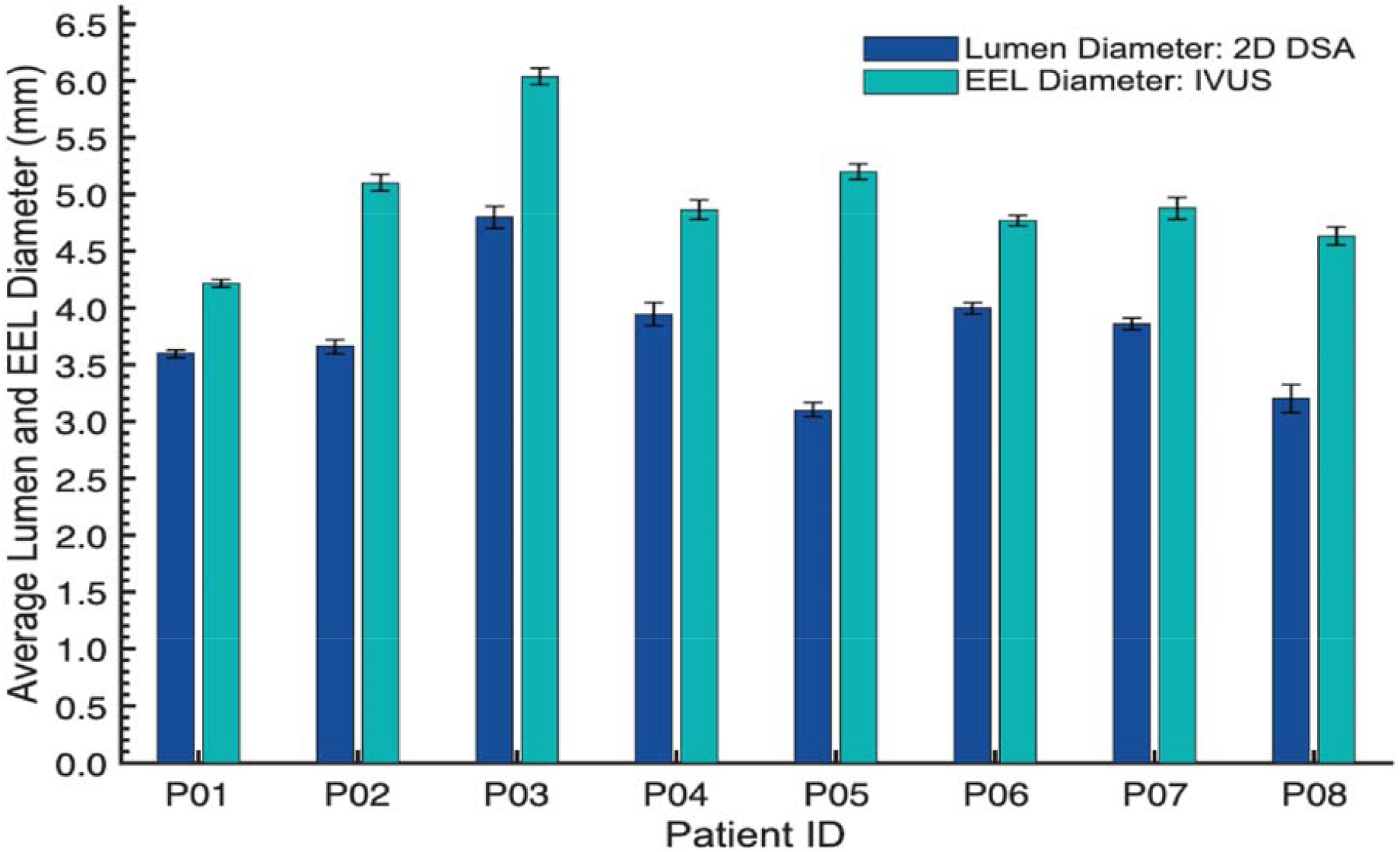
Comparing Lumen and External Elastic Lamina (EEL) Diameters. Mean lumen and EEL diameter ± standard error from eight patients with stenotic vertebral artery origins. The mean percent change from lumen to EEL diameter was 32.96%, with a mean difference of 1.19 mm between lumen and EEL diameter measurements.

Figure 3 presents all lumen and EEL measurements along each patient’s vertebral artery. Wilcoxon signed-rank tests were used to compare lumen to EEL diameters (p<0.05). It should be noted that since this study investigates two anatomically distinct measurements, agreement statistics were not used.

**Figure 3.**
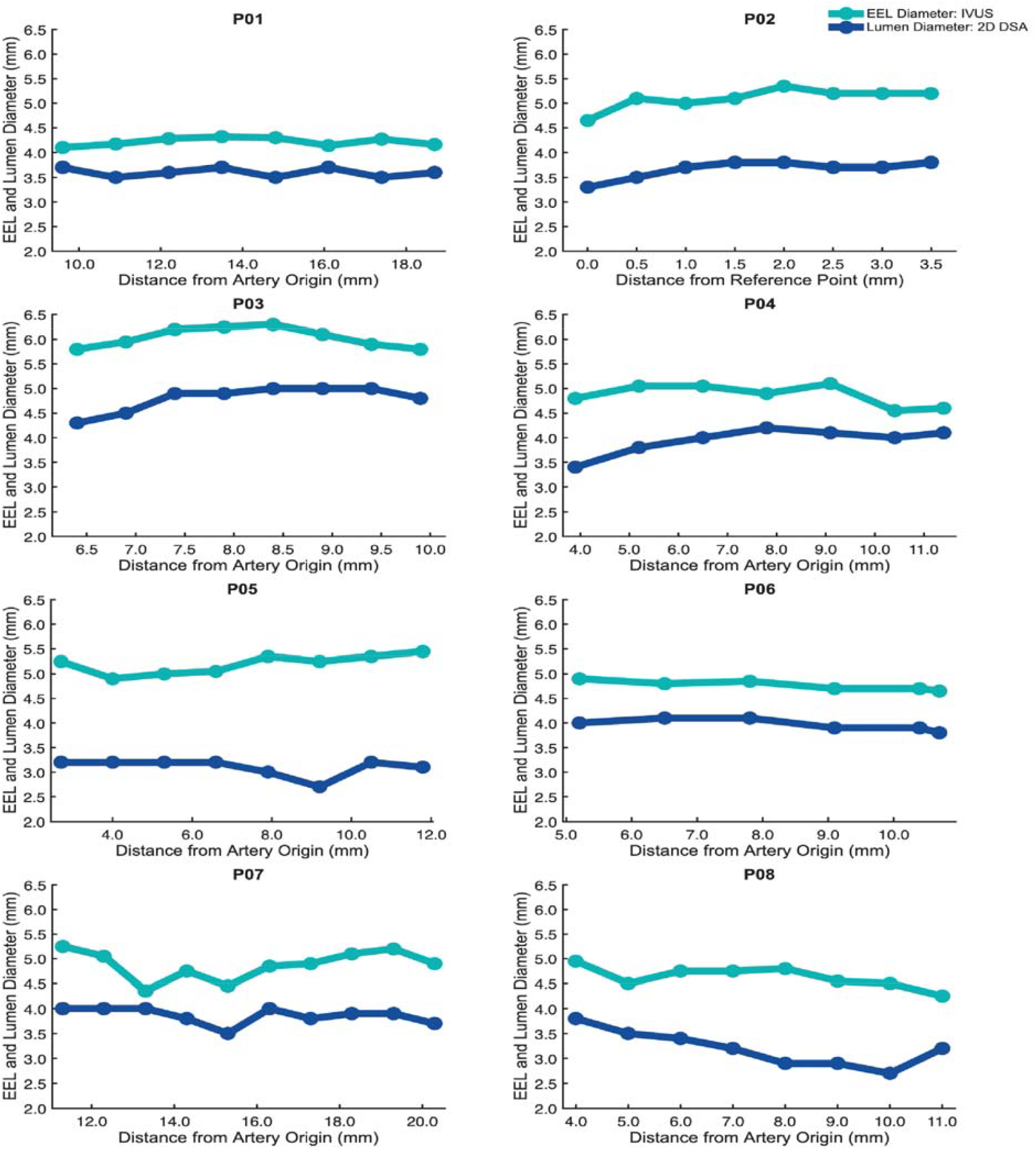
Comparing External Elastic Lamina (EEL) to Lumen Vertebral Artery Diameters Across Patients. EEL and lumen diameter were measured at each point of interest along the vertebral artery for all patients. While 2-dimensional digital subtraction angiography can only measure the lumen diameter of arteries, intravascular ultrasound can measure the EEL diameter of the arterial walls. Wilcoxon signed-rank tests were used to compare the lumen to EEL diameters in each patient (p<0.05). Patient P02’s measurements were not taken relative to the vertebral artery origin due to a high degree of stenosis and tortuosity.

## Conclusions and Discussion

The data demonstrate a statistically significant difference between vessel size measurements obtained from 2D DSA as compared to the more precise IVUS measurements. This deviation suggests that the utilization of IVUS may be very valuable, particularly in critical situations such as sizing stents, where accuracy is of paramount importance, and undersizing can pose life-threatening consequences.

2D DSA is restricted to measuring only the arterial luminal diameter,[3] as the contrast can only be imaged as contained by the tunica intima. Consequently, the technique is unable to measure the EEL diameter. Thus, 2D DSA is limited in its ability to provide estimates for sizes of arterial stents, and measurements derived from this technique can lead to undersizing stents. This increases the risk of restenosis or, in more severe cases, stent thrombosis and/or stent movement. Consistently and significantly larger measurements were obtained from IVUS due to its ability to measure the EEL.

### Windowing Limitations

Using constant window parameters for 2D DSA lumen measurements ensured a reliable comparison between patients, but it introduced some difficulty in visualizing contrast within the vertebral artery. To address this issue, other windows were used to corroborate the boundaries of the arterial lumen. Nevertheless, lumen diameters were measured using a constant window for all patients. Future studies may benefit from characterizing how windowing affects vessel diameter measurements. Any error introduced by windowing was likely negligible and included in the standard error.

### A Note on IVUS

For IVUS, as opposed to angiography, the imaging catheter must pass the lesion to record images. This can increase the risk of vessel damage, including dissection and occlusion. In contrast, angiograms require the catheter to be placed proximal to the lesion, and images can be obtained without crossing the lesion. The use of IVUS also significantly decreases contrast use,[7] and while increased procedure time from IVUS is a disadvantage, it may be limited by proper training.[7]

Further, research on percutaneous coronary intervention (PCI) has highlighted the economic advantage of IVUS, with 99% of Monte Carlo iterations run concluding IVUS as cost-effective compared to angiography.[8] Analysis of conservatively estimated incremental cost-effectiveness ratios supported the same conclusion, with each additional quality-adjusted life year (QALY) valued at $26,601.[8]

### Future Studies

The cases studied focused exclusively on vertebral arteries. Future studies should examine the difference between IVUS and 2D DSA within additional vascular territories. Additionally, we did not study other anatomical lesions that may need stents, such as aneurysms.

We also compared IVUS to 3D rotational digital subtraction angiography using flow models. Such data were excluded from the study as both the plastic-based PETG and silicone flow models used could not model the surrounding tissue. PVA-cryogel may be a viable solution to this, as varying the number of freeze-thaw cycles in the gel can alter its mechanical and acoustic characteristics, and when paired with proper scattering agents, can improve ultrasound image contrast.[9] Future studies can evaluate models with vessels that allow for smooth movement of catheters and IVUS probes and distinctly display the vessel wall.[9]

Future analysis might also benefit from investigating vessel foreshortening introduced by changing rostral and caudal angles on 2D DSA.

## Data Availability

The datasets generated and analyzed during the current study are available from the corresponding author upon reasonable request.

